# Establishing an empirical cut-off on the 12-item Brief Berger HIV Stigma Scale to screen psychosocial vulnerability among PLHIV in Nigeria

**DOI:** 10.1101/2025.09.15.25335818

**Authors:** Kamaldeen Sunkanmi Abdulraheem, Adebayo T. Onajole, Alero Ann Roberts, Temidayo Olowoopejo

## Abstract

This study addresses a key challenge in HIV care the lack of a validated screening tool for psychosocial vulnerability. Although the 12-item Brief Berger HIV Stigma Scale is widely used, no clear threshold exists to identify individuals at high risk for mental health problems. Our research aimed to establish a practical, data-driven cut-off score for the scale in a Nigerian context and to explore which dimensions of stigma are most linked to psychosocial vulnerability.

We conducted a cross-sectional study of 285 PLWH at three tertiary centres (May–August 2024). Psychosocial vulnerability was defined as moderate-to-severe depression (PHQ-9 ≥10) or anxiety (GAD-7 ≥10). The Receiver Operating Characteristic (ROC) analysis with the Youden Index identified the optimal cut-off; multivariable logistic regression examined independent associations of stigma subscales with vulnerability.

Among 285 participants (mean age 47.1±11.17 years, 72.6% female), 44.9% met vulnerability criteria. The Berger Scale demonstrated acceptable discrimination (AUC = 0.717, 95% CI 0.658–0.777). A cut-off of ≥30 yielded high sensitivity (87.5%) and strong negative predictive value (82.8%). Internal validation confirmed stability (cross-validated

AUC 0.703, bootstrap AUC 0.701). Decision curve analysis showed positive net benefit over “screen-none” up to threshold probability 0.45, with peak benefit at 0.30. In multivariable analysis, public attitude concerns were the strongest predictor (adjusted OR 1.68, p<0.001), while disclosure concerns—despite near-universal prevalence (96.1%)—showed no independent association.

The ≥30 cut-off provides a practical, sensitive rule-out tool for identifying PLWH needing psychosocial assessment in resource-limited settings. **External validation is essential before widespread adoption**. Public attitude concerns outweighing internalised stigma highlights the need for culturally informed interventions addressing societal stigma alongside mental health support.

## Introduction

Human Immunodeficiency Virus (HIV) remains a global public health challenge, disproportionately affecting sub-Saharan Africa (SSA), which accounts for over 67% of the disease burden.^1^ In 2021, SSA accounted for 670,000 of the 1.5 million new infections and 280,000 of the 650,000 AIDS-related mortality worldwide.^1^ There are over 1.9 million people living with HIV (PLHIV) in Nigeria, with a prevalence of 1.4%, making it the second largest epidemic globally.^2^

Psychosocial vulnerability is an individual’s greater susceptibility to mental and physical health problems caused by the interaction of psychological and social elements, as well as stressful life experiences.^3^ The psychological toll of HIV is substantial. A Systematic review has reported that depression affects 20-36% of PLHIV in Africa, while anxiety prevalence among young people in SSA can range from 2.2% to 25.0%.^4,5^ An estimated 28.2% of PLHIV in Nigeria experiencing major depression over a 12-month period.^6^

One of the critical psychosocial dimensions of HIV is the disclosure of one’s serostatus. Deciding whether to disclose one’s HIV status is a complex process shaped by cultural norms, relationship dynamics, fear of stigma, and anticipated reactions from others.^7^ HIV-related stigma and discrimination remain pervasive and exert profound effects on the mental health and well-being of PLHIV, creating major barriers to care engagement and adherence to antiretroviral therapy.^8,9^ Fear of enacted stigma which include verbal abuse, social isolation, loss of employment, and family rejection — is the primary driver of non-disclosure in SSA and consistently exacerbates depression and anxiety.^10–12^

HIV-related stigma is a complex construct with multiple dimensions, including personalised stigma, disclosure concerns, negative self-image, and anxieties about public attitudes.^13^ This stigma is consistently linked to psychosocial distress and nondisclosure of HIV status.^14^ A failure to disclose one’s status not only worsens an individual’s own health outcomes, such as poor viral suppression and increased morbidity, but also contributes to the ongoing transmission of the virus.^15^ This interconnectedness highlights why psychosocial vulnerability, often manifesting as depression and anxiety, is so strongly linked to stigma and nondisclosure.^16^

Despite the widespread use of the 12-item Brief Berger HIV Stigma Scale to measure these issues, a practical threshold to identify individuals at the greatest psychological risk remains a critical gap in research.^17^ In Nigeria, where HIV prevalence is substantial and stigma poses significant challenges to effective disease management, the absence of validated cut-off scores limits the clinical utility of stigma assessment tools.^2,18^ This represents a missed opportunity for early intervention and targeted mental health support in a high-prevalence setting.

This study was therefore designed to address this deficiency. We aimed to establish an empirically derived cut-off score for the 12-item Brief Berger HIV Stigma Scale that can predict psychosocial vulnerability among PLHIV in Nigeria. By using receiver operating characteristic (ROC) analysis, we sought to determine an optimal threshold that effectively balances sensitivity and specificity.^19^ Additionally, we examined which specific dimensions of HIV stigma most strongly predict psychosocial vulnerability. The findings from this research will help clinicians develop more targeted interventions and enhance integrated HIV care.

## Methods and Materials

### Ethical Approval

Ethical approval was obtained from the institutional review boards of all participating institutions LUTH Health Research Ethics Committee (HREC assigned no: ADM/DCST/HREC/APP/6242) and NIMR Institutional Review Board (IRB/24/057). Administrative approval was also obtained from the Lagos State Ministry of Health (LSMH/4682/l/126) for LASUTH. The study was conducted in accordance with the ethical principles outlined in the **Declaration of Helsinki**. All participants provided written informed consent before enrolment, after a thorough explanation of the study’s purpose, procedures, and their right to withdraw at any time. Participant confidentiality was maintained using unique study identifiers. All data were stored securely in a password-protected computer with restricted access, ensuring that no personally identifiable information was linked to the responses.

### Study Location

This study was conducted in Lagos State, Nigeria, located in the southwestern region of the country. With an estimated population of over 15 million by 2022, Lagos is the most populous city in Africa.^20^ The study was conducted in three tertiary centres that provide HIV care in Lagos State: Lagos University Teaching Hospital (LUTH), Lagos State University Teaching Hospital (LASUTH), and the Nigerian Institute of Medical Research (NIMR). LUTH and NIMR are PEPFAR-supported and offer comprehensive HIV services. LASUTH, a major referral hospital, operates its Haematology Clinic four days a week, attending to approximately 100 patients daily.

### Study Design

This was an analytical cross-sectional study aimed at exploring HIV serostatus disclosure and establishing an empirical cut-off score for the 12-item Brief Berger HIV Stigma Scale to identify psychosocial vulnerability among PLHIV.

### Eligibility Criteria

Participants were required to be 18 years or older, diagnosed with HIV/AIDS, receiving care at one of the participating tertiary healthcare centres in Lagos, and in stable medical condition as determined by their attending physician. Individuals were excluded, however, if they were unable to provide informed consent due to severe illness or mental incapacitation.

### Sample Size Estimation

The initial minimum sample size was determined using the Cochran formula^21^ for proportions to ensure adequate precision for estimating secondary descriptive parameters, such as HIV serostatus disclosure prevalence. Using an expected disclosure prevalence of 81.9% (based on prior literature)^22^, a 95% confidence level, and 5% precision (d = 0.05), a minimum sample of 228 participants was calculated.

The final target sample size was conservatively set at 285 to account for potential non-response (estimated at 20%) and to ensure sufficient outcome-positive and outcome-negative cases for the primary objective: diagnostic accuracy of the Brief Berger Scale in predicting psychosocial vulnerability. This sample size ensured stable discrimination estimates in ROC analysis and adequate events-per-variable ratio (≥10) for multivariable logistic regression modelling.

Post-hoc assessment confirmed adequate precision of the diagnostic accuracy findings.^23^ With 128 positive and 157 negative cases for the composite psychosocial vulnerability outcome, the observed AUC of 0.717 was estimated with a standard error of 0.031, yielding a 95% CI width of 0.12 (95% CI 0.658–0.777). This narrow confidence interval supported the reliability and stability of the diagnostic accuracy estimates.

### Sampling Methodology

Proportionate stratified sampling was employed based on clinic patient load: 106 participants from LUTH, 86 from LASUTH, and 93 from NIMR. On each clinic day, a sampling frame was created from scheduled patient card numbers, and simple random sampling by balloting was used to select participants until the site-specific quota was reached. Replacement sampling was performed for those who declined or were ineligible. A STROBE-compliant participant flow diagram is provided in Figure 1. No demographic comparisons were available for non-participants.

**Figure 1.**
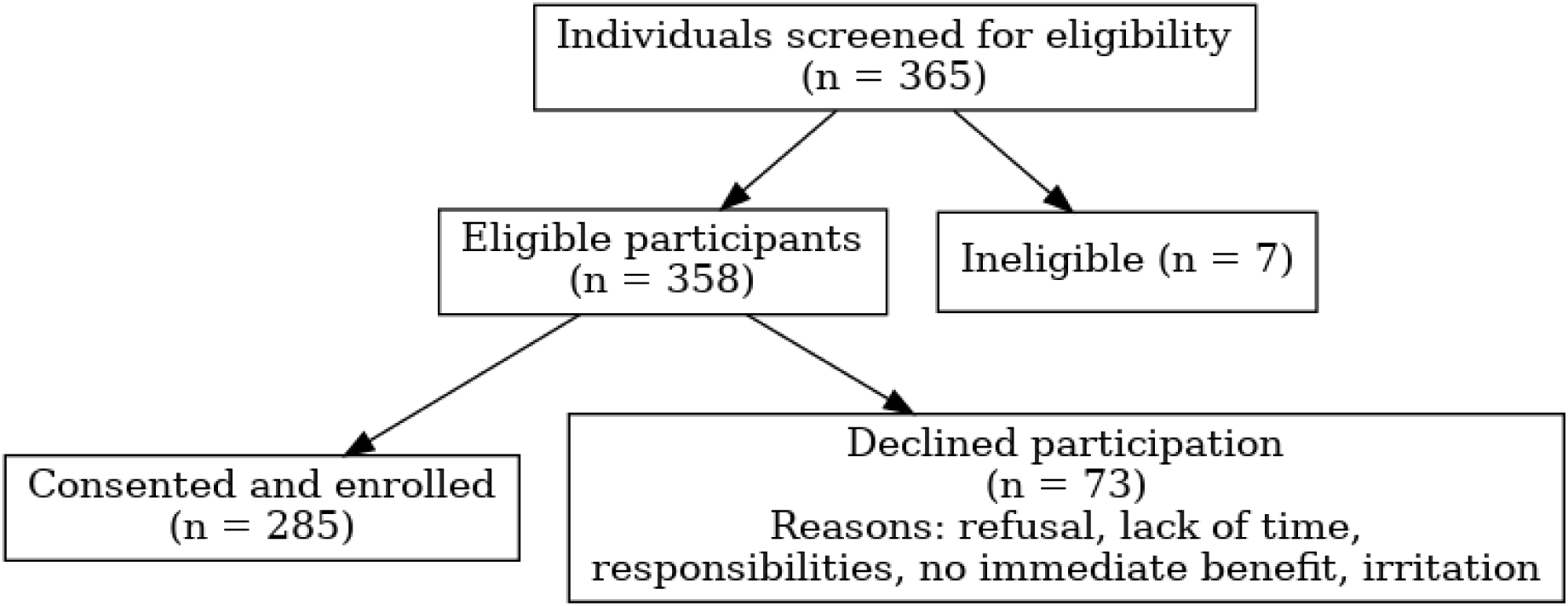
Participant Flow Chart.

### Data Collection Tool

The 12-item Brief Berger HIV Stigma Scale which is divided into four subscales (personalised stigma, negative self-image, disclosure concerns, and public attitudes) was used assess HIV-related stigma. We used the 12-item Brief Berger HIV Stigma Scale because it has been validated across diverse settings, retains the four core dimensions of the original 40-item scale, and demonstrates strong reliability. The brief version substantially reduces respondent burden in busy ART clinics, making it more feasible for routine screening—consistent with our goal of deriving a clinically practical cut-off. The cumulative score for all items on the brief Berger scale fell within the range of 12 to 48. Depression was assessed using the Patient Health Questionnaire-9 (PHQ-9). The total score on the PHQ-9 ranged from 0 to 27. Anxiety was assessed using the Generalized Anxiety Disorder-7 (GAD-7) scales with total scores ranging from 0 to 21. All tools were administered in English, the primary language of clinical care in Lagos. Linguistic appropriateness was confirmed during pretesting with 30 PLHIV, and these scales have been previously used within Nigerian PLHIV populations.^17^

### Variables

#### Definition of Psychosocial Vulnerability

Psychosocial vulnerability was defined as a composite dichotomous outcome whereby participants were classified as vulnerable if they scored ≥10 on the PHQ-9; indicating moderate-to-severe depression or ≥10 on the GAD-7; indicating moderate-to-severe anxiety, with all others classified as not vulnerable. This threshold was selected as it corresponds to the clinically recommended level for initiating treatment or referral in primary care settings. The composite approach was selected to increase screening sensitivity and better reflect real-world clinical patterns in which depression and anxiety frequently co-occur among people living with HIV, thereby facilitating early identification of individuals who may benefit from psychosocial support.

The following independent variables were included in the analysis: sociodemographic variables such as age group (21–30, 31–40, 41–50, and ≥51 years), gender (male and female), level of education (no formal education, primary, secondary, and tertiary), marital status (married, single, separated, divorced, widowed, and co-habiting), average monthly income (less than =50,000, =51,000–=99,000, and greater than =100,000), and employment status (full-time, part-time, retired, and unemployed). These variables were categorized based on our study protocol and are consistent with standard demographic reporting in public health research. The categorical definitions were chosen to align with existing research to facilitate meaningful comparisons with other national health studies. The primary independent variable was the HIV stigma score, as assessed by the 12-item Brief HIV Stigma Scale.

#### Data Collection Procedure

Data collection was conducted by trained research assistants who administered pretested questionnaires to ensure accuracy and consistency. Data collection took place over four months (May to August 2024) and continued until the required sample size was achieved.

Recruitment did not begin simultaneously across sites: participants were recruited at LUTH from 15 May to 14 June 2024, at LASUTH from 1 to 26 July 2024, and at NIMR from 15 to 30 August 2024. Where necessary, two medical doctors or trained research assistants provided interviewer-administered support and read questions to participants who were not literate in English. To ensure data quality and minimize participant burden, all questionnaires were inspected for completeness immediately upon collection, which resulted in no missing data for key analytical variables.

#### Quality Assurance

Questionnaires were pretested on a small sample (30 PLHIV, approximately 10%) to ensure clarity and cultural appropriateness. Research assistants received supportive supervision.

#### Statistical Analysis

All data were cleaned in Microsoft Excel and analysed using R (Version 2024.12.0+467). Descriptive statistics were computed to summarize participants’ socio-demographic characteristics, HIV-related stigma scores, and mental health indicators (depression and anxiety). The analysed data were presented using frequency tables and charts to provide a clear and accessible overview of the findings. Categorical variables were summarised using frequencies and proportions, while continuous variables were presented as means with standard deviations (SD), after assessing normality through visual inspection of histograms and the Shapiro–Wilk test (p ≥ 0.05 indicating normality).

The internal consistency of the 12-item Berger HIV Stigma Scale was evaluated using Cronbach’s alpha, with a reliability coefficient above 0.70 considered satisfactory.^18^ To establish an empirical cut-off, point for the 12-item Berger HIV Stigma Scale in identifying psychosocial vulnerability, stigma scores were treated as a continuous variable. A Receiver Operating Characteristic (ROC) curve analysis was performed to determine the sensitivity and specificity of different stigma score thresholds in predicting psychosocial vulnerability. The Youden Index was used to determine the optimal cut-off score, maximizing the balance between sensitivity and specificity. The optimal cut-off was identified using the Youden Index (maximising sensitivity + specificity – 1). The Area under the curve (AUC), sensitivity, specificity, positive and negative predictive values, with 95% Wilson confidence intervals, were reported. An AUC value of 0.7 or greater considered to have acceptable discriminatory power, as is standard by Hosmer-Lemeshow.^19^ Decision curve analysis was conducted to assess clinical net benefit across threshold probabilities of 20–60%.

Multivariable logistic regression was used to assess the independent associations of the four Berger HIV Stigma subscales (personalised stigma, negative self-image, disclosure concerns, and concerns with public attitudes) with psychosocial vulnerability. Age (treated as a continuous variable), sex, and marital status were forced a priori into every model because they are established sociodemographic determinants of both HIV-related stigma and mental health outcomes among people living with HIV in sub-Saharan Africa and are routinely adjusted for in comparable Nigerian studies. All other candidate variables with p < 0.20 in bivariate analysis were initially entered together with the four stigma subscales. Backward elimination was then performed using the Akaike Information Criterion (AIC) as the primary guide; at each step, the variable whose removal produced the largest decrease in AIC was dropped. Throughout the process, events-per-variable ratio (EPV) and multicollinearity were actively monitored to prevent overfitting and instability — no model with EPV < 10 or any variance inflation factor (VIF) > 5 was accepted. Although several stigma subscales, including disclosure concerns, were statistically significant in bivariate analysis, they were removed during backward elimination because their inclusion increased the AIC, reduced the EPV (to as low as 12.6 in intermediate models), and raised VIF values above 2 for the remaining subscales without substantially altering the magnitude or significance of the coefficients of the retained subscales (change < 10%). The final parsimonious model therefore retained only the concerns with public attitudes and negative self-image subscales alongside the three forced sociodemographic variables. This model achieved the lowest AIC (351.4), excellent discrimination (AUC 0.763, 95% CI 0.709–0.817), good calibration (Hosmer-Lemeshow p = 0.43), no multicollinearity (maximum VIF 1.34), and a comfortable EPV of 14.7, meeting recommended thresholds for stability and generalisability.

#### Robustness and Generalisability Assessments

To assess the robustness and generalisability of the proposed cut-off score for the 12-item Brief Berger HIV Stigma Scale, a series of complementary analyses were conducted.

First, sensitivity analyses were performed by repeating the ROC curve analysis with modified definitions of the outcome. These included using moderate-to-severe depression alone (PHQ-9 ≥10), moderate-to-severe anxiety alone (GAD-7 ≥10), and stricter clinical thresholds for both instruments (PHQ-9 ≥15 and GAD-7 ≥15 indicating severe symptoms). These modifications tested whether the optimal stigma cut-off and overall discriminatory performance remained stable when the composite vulnerability definition was altered.

Second, subgroup analyses were conducted to examine whether the diagnostic performance of the stigma scale differed by sex. Separate ROC curves were generated for male and female participants, and area under the curve (AUC) values were compared descriptively.

Third, to determine whether any observed differences in subgroup performance were statistically significant (i.e., evidence of effect modification or interaction by sex), formal testing for interaction was performed using DeLong’s test to compare the AUCs between males and females.

Results of these analyses showed consistent discriminatory performance across alternative outcome definitions (AUC range 0.717–0.750), similar AUC values in men and women, and no evidence of statistically significant interaction (DeLong’s test p > 0.8). Additional calculations of positive and negative predictive values at hypothetical lower prevalence levels (10% and 25%) were performed to facilitate application of the cut-off in settings with different baseline rates of psychosocial vulnerability (see Supplementary tables). Clinical utility across a range of threshold probabilities was further evaluated using decision curve analysis.

## RESULT

A total of 365 individuals were screened for eligibility across the three study sites. Of these, 358 met the inclusion criteria. The final sample comprised 285 participants, as 73 individuals declined to participate. The main reasons for non-participation included refusal, lack of time, the need to collect medications or attend to other responsibilities, absence of immediate monetary benefit, and perceived disturbance or irritation.

The study included 285 participants with a mean age of 47.1 years (SD ± 11.17), predominantly in the 41-50 age group (43.51%) and over 50 years (34.04%). The sample was predominantly female (72.63%) and married (65.61%), with most participants having secondary (41.05%) or tertiary (39.65%) education. Economically, half the participants earned less than 50,000 naira monthly (50.88%), with the majority employed part-time (54.74%) and about a quarter in full-time employment (23.51%). Only 17.19% were unemployed. This demographic profile represents a predominantly middle-aged, educated, urban population engaged with formal HIV care services at tertiary healthcare facilities in Lagos.

**Table 1:** Sociodemographic characteristics of respondents.

| **Variables** | **Frequency (n= 285)** | **Percentage (%)** |
| --- | --- | --- |
| **Age group (years)** |  |  |
| 21-30 | 25 | 8.77 |
| 31-40 | 39 | 13.68 |
| 41-50 | 124 | 43.51 |
| $\geq 51$ | 97 | 34.04 |
| **Mean age (SD)** | 47.12 ± 11.17 | |
| **Gender** |  |  |
| Female | 207 | 72.63 |
| Male | 78 | 27.37 |
| **Level of Education** |  |  |
| No formal education | 23 | 8.07 |
| Primary | 32 | 11.23 |
| Secondary | 117 | 41.05 |
| Tertiary | 113 | 39.65 |
| **Marital Status** |  |  |
| Married | 187 | 65.61 |
| Single | 38 | 13.33 |
| Separated | 34 | 11.93 |
| Divorced | 14 | 4.91 |
| Widow | 7 | 2.46 |
| Co-habiting | 5 | 1.75 |
| **Average monthly Income** |  |  |
| Less than 50,000 | 145 | 50.88 |
| 51,000- 99, 000 | 106 | 37.19 |
| ≥ 100,000 | 34 | 11.93 |
| **Employment Status** |  |  |
| Full-time | 67 | 23.51 |
| Part-time | 156 | 54.74 |
| Retired | 13 | 4.56 |
| Unemployed | 49 | 17.19 |
| **Total** | **285** | **100.0** |

*[Place Figure 2. Here]* **Figure 2: Pie Chart showing the prevalence of HIV disclosure**

**Figure 2:**
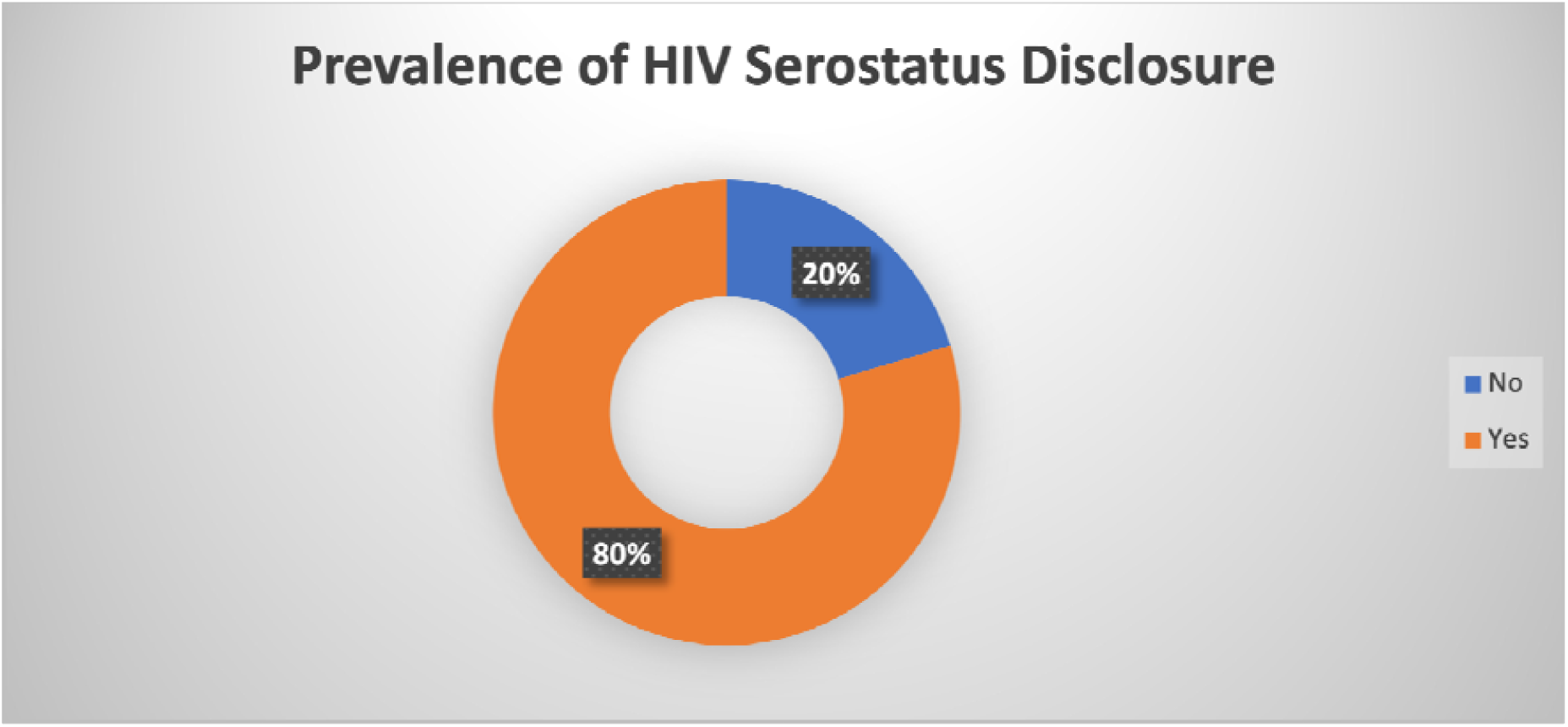
Pie Chart showing the prevalence of HIV disclosure serostatus.

Majority of the respondents have disclosed their HIV status 80% while 20% have not disclosed their HIV status.

### Diagnostic Accuracy

Receiver operating characteristic (ROC) curve analysis revealed acceptable discriminatory performance of the total stigma score for detecting psychosocial vulnerability (area under the curve [AUC] = 0.717, 95% CI 0.658–0.777).

**Figure 3:**
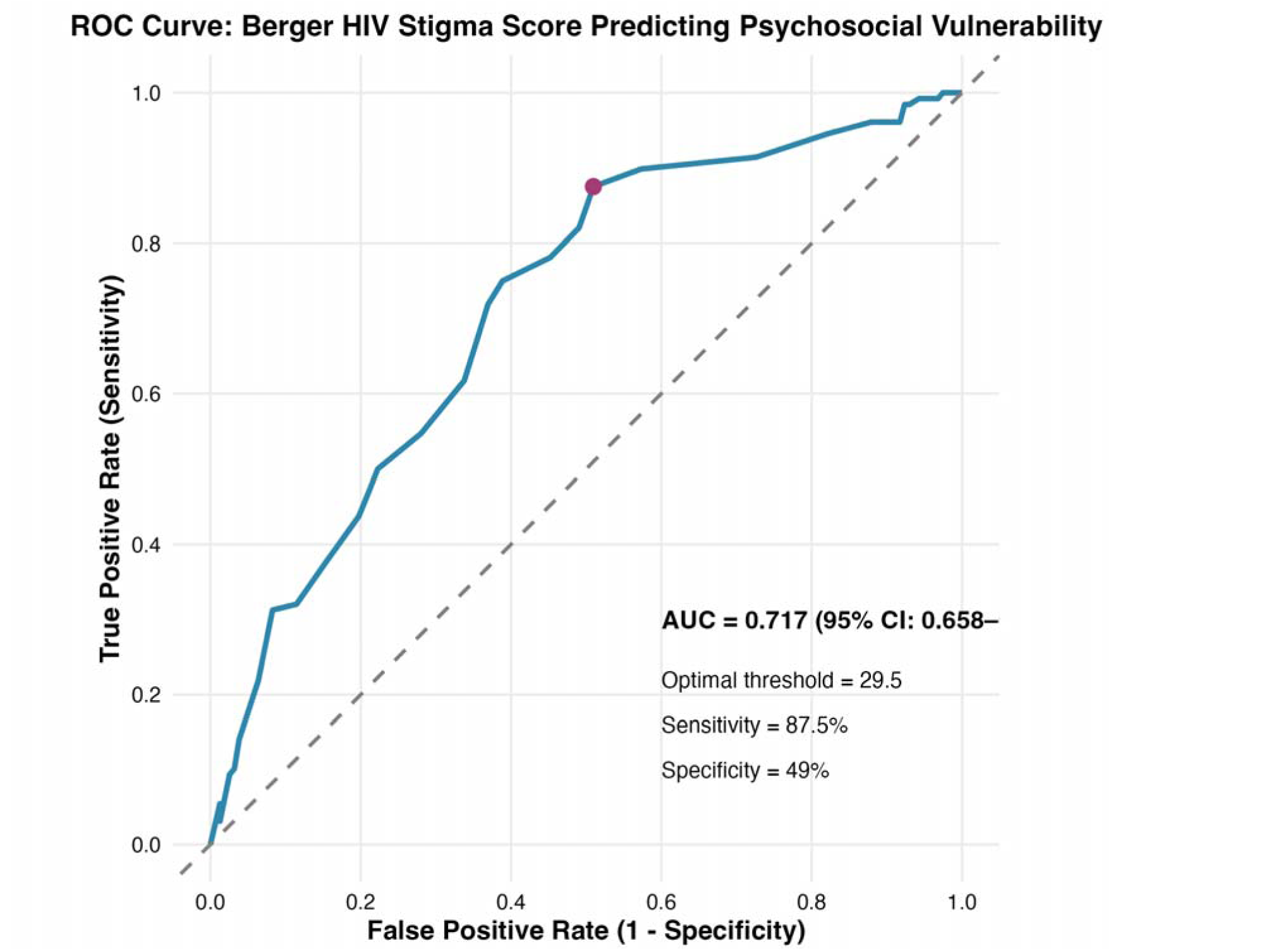
Receiver operating characteristic curve for the Berger HIV Stigma Score predicting psychosocial vulnerability.

At the clinically practical cut-off of ≥30, the 12-item Brief Berger HIV Stigma Scale achieved high sensitivity (87.5%, 95% CI 80.6–92.2%) and strong negative predictive value (82.8%, 95% CI 73.9–89.1%).

**Table 2.** Diagnostic accuracy metrics of the 12-item Brief Berger HIV Stigma Scale using the clinically recommended cut-off of≥30.

| **Metric at cut-off $\geq 30$** | **Value** | **95% CI (Wilson)** |
| --- | --- | --- |
| Sensitivity | 87.5% | 80.6% – 92.2% |
| Specificity | 49.0% | 41.4% – 56.7% |
| Positive predictive value (PPV) | 58.3% | 51.1% – 65.2% |
| Negative predictive value (NPV) | 82.8% | 73.9% – 89.1% |

Internal validation using 10-fold cross-validation and 1000 bootstrap resamples confirmed minimal overfitting, with an optimism-corrected AUC of 0.701 (compared with apparent AUC 0.717), supporting the robustness of the stigma scale as a screening tool.

**Table 3.**
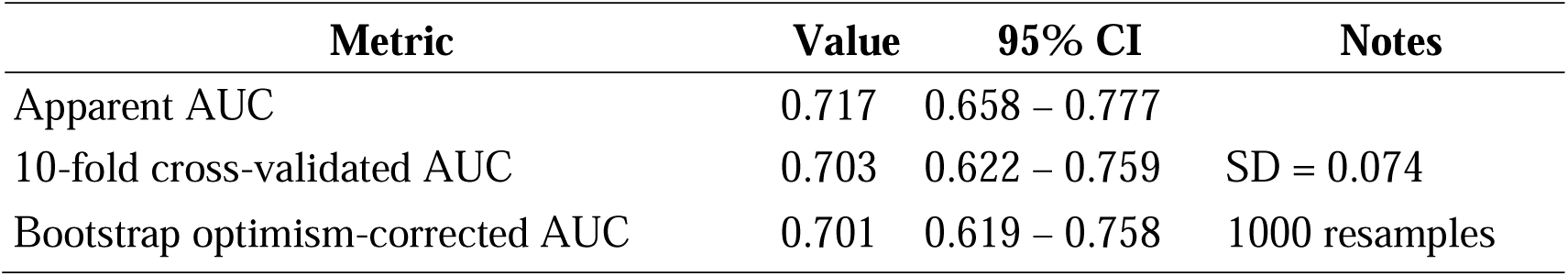
Internal validation of the discriminatory performance of the 12-item Brief Berger HIV Stigma Scale total score for detecting psychosocial vulnerability.

| Metric | Value | 95% CI | Notes |
| --- | --- | --- | --- |
| Apparent AUC | 0.717 | 0.658 – 0.777 |  |
| 10-fold cross-validated AUC | 0.703 | 0.622 – 0.759 | SD = 0.074 |
| Bootstrap optimism-corrected AUC | 0.701 | 0.619 – 0.758 | 1000 resamples |

*[Place Figure 4 here]* **Figure 4: Decision Curve Analysis (DCA) for the Stigma Score Cut-off ( $\geq 30$ ) in Predicting Psychosocial Vulnerability.**

**Figure 4:**
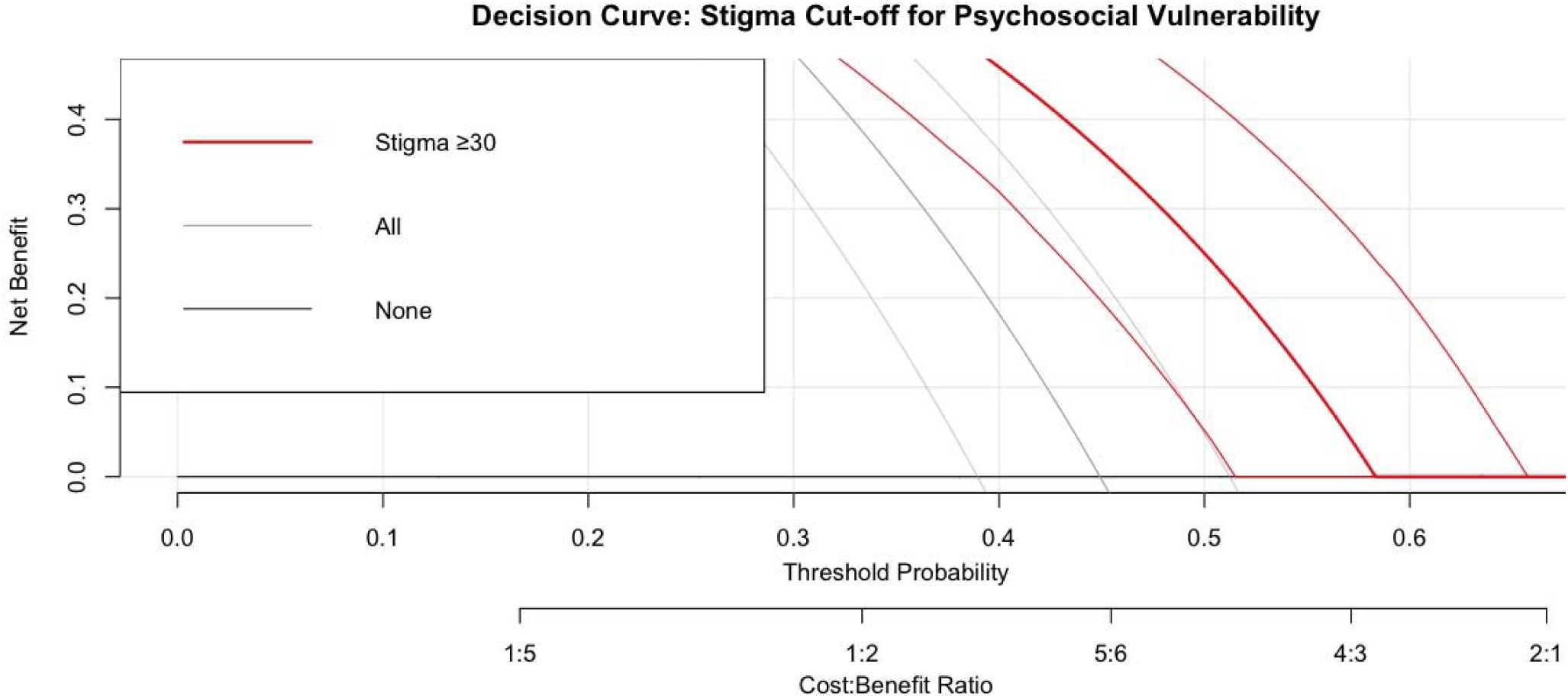
Decision Curve Analysis (DCA) for the Stigma Score Cut-off (≥30) in Predicting Psychosocial Vulnerability.

The ≥30 cut-off yielded positive net benefit over the ‘treat none’ strategy across clinically relevant threshold probabilities, with peak benefit around a threshold of 0.30.

The model yielded positive net benefit relative to “treat none” up to a threshold probability of **0.45**, with peak benefit at **threshold = 0.30** (NB = 0.27). Net benefit declined progressively at higher thresholds (NB = 0.21 at 0.40; 0.16 at 0.45; 0.11 at 0.50), and approached zero beyond **0.55**, indicating that the model is clinically useful only within the lower decision-threshold range.

### Sensitivity and Subgroup Analyses

The prevalence of psychosocial vulnerability was significantly higher among female participants (49.8%) than male participants (32.1%; χ² p = 0.004). Discriminatory performance was comparable across sexes. Formal testing with DeLong’s test revealed no significant interaction by sex (p = 0.895)

**Table 4:** Subgroup analysis of diagnostic performance of the 12-item Brief Berger HIV Stigma Scale by sex.

| Metric | Females (n=183) | Males (n=102) | Comparison p-value |
| --- | --- | --- | --- |
| Prevalence | 49.8% | 32.1% | 0.004 ($\chi^2$) |
| AUC (95% CI) | 0.716 (0.640 – 0.792) | 0.706 (0.604 – 0.808) | 0.895 (DeLong) |
| Optimal Youden cut-off | $\geq 29.5$ | $\geq 33.0$ | — |
| Sensitivity at own cut-off | 89.3% | 72.0% | — |
| Specificity at own cut-off | 48.1% | 60.4% | — |
| PPV at own cut-off | 58.1% | 42.9% | — |
| NPV at own cut-off | 82.0% | 82.1% | — |

The 12-item Brief Berger HIV Stigma Scale retained acceptable discrimination (AUC ≥0.720) and consistently high sensitivity and negative predictive value across definitions based on depression alone, anxiety alone, or stricter clinical thresholds

**Table 5.** Sensitivity analyses of the diagnostic performance of the 12-item Brief Berger HIV Stigma Scale using alternative definitions of psychosocial vulnerability.

| Outcome Definition | Prevalence (n, %) | AUC (95% CI) | Optimal Threshold (Youden) | Sensitivity | Specificity | NPV |
| --- | --- | --- | --- | --- | --- | --- |
| PHQ-9 $\geq 10$<br>(moderate–severe depression) | 107 (37.5%) | 0.730<br>(0.670 – 0.790) | $\geq 32.5$ | 79.4% | 59.6% | 83.2% |
| PHQ-9 $\geq 15$ (severe depression) | 64 (22.5%) | 0.750<br>(0.680 – 0.820) | $\geq 33.5$ | 85.9% | 57.0% | 93.4% |
| GAD-7 $\geq 10$<br>(moderate–severe anxiety) | 99 (34.7%) | 0.720<br>(0.660 – 0.780) | $\geq 29.5$ | 91.9% | 45.7% | 90.9% |
| GAD-7 $\geq 15$ (severe anxiety) | 26 (9.1%) | 0.720<br>(0.620 – 0.810) | $\geq 30.5$ | 96.2% | 39.4% | 99.0% |

In multivariable logistic regression, higher scores on the Concerns with Public Attitudes subscale were independently associated with greater odds of psychosocial vulnerability (adjusted OR 1.68 per 1-point increase, 95% CI 1.34–2.14, p < 0.001). The Negative Self-Image subscale showed a trend toward significance (adjusted OR 1.36, 95% CI 0.98–1.89, p = 0.066). Male sex was protective (adjusted OR 0.52, 95% CI 0.28–0.95, p = 0.035). Age, employment status, and marital status were not statistically significant after adjustment.

**Table 6:**
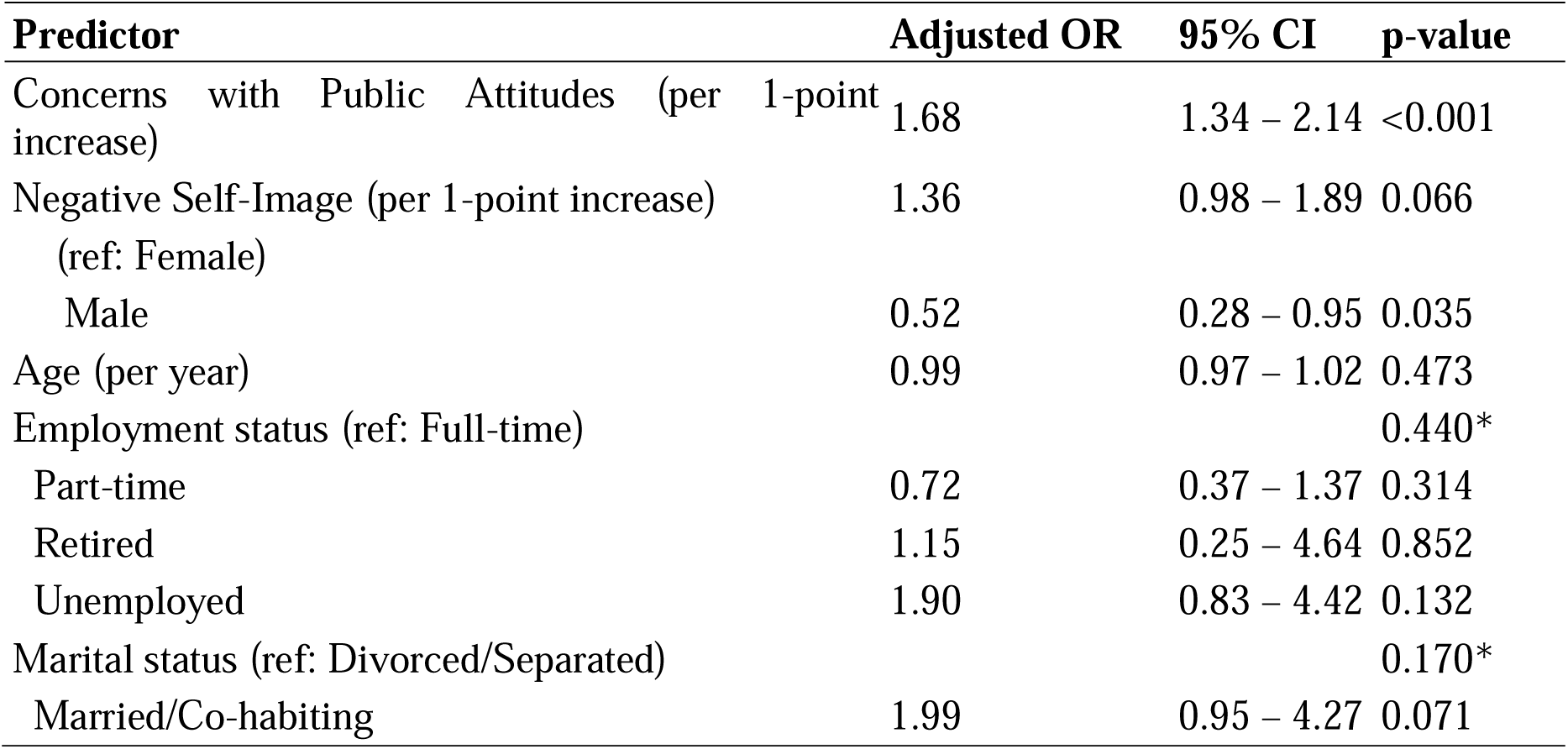
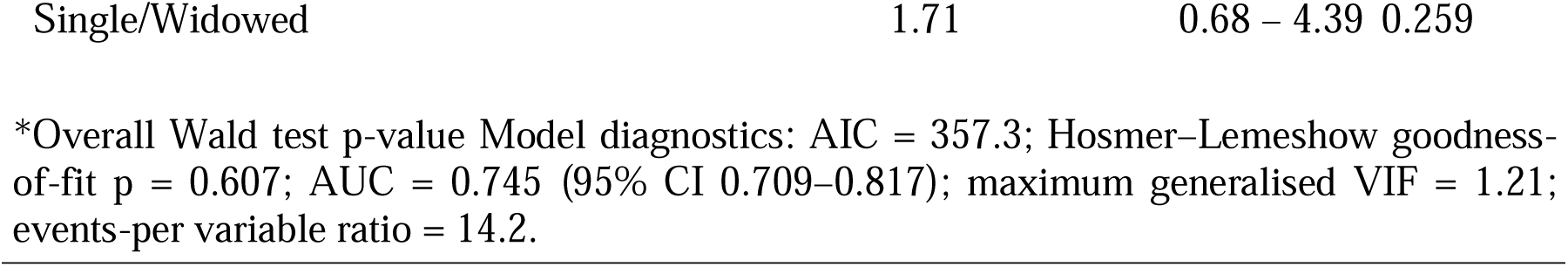
Multivariable logistic regression model predicting psychosocial vulnerability (n = 285, 128 events)

## DISCUSSION

### Principal Findings

This study validated the 12-item Brief Berger HIV Stigma Scale as a screening tool for identifying psychosocial vulnerability (depression or anxiety) among people living with HIV in tertiary facilities in Lagos, Nigeria. The scale achieved acceptable discriminatory performance (AUC = 0.717), with an operational cut-off of ≥30 yielding high sensitivity (87.5%) and strong negative predictive value (82.8%). Only 20% had not disclosed their HIV status to at least one person. Concerns about public attitudes emerged as the strongest predictor of psychosocial vulnerability, while internalised stigma showed weaker associations than expected.

### Diagnostic Accuracy and Clinical Utility

An AUC of 0.717 is at the low end of acceptable discrimination which is considered adequate discrimination for screening purposes.^24^ The modest specificity (49%) is acceptable for a high-sensitivity screening tool and indicates that positive screens require confirmatory assessment (PHQ-9/GAD-7). This may increase workload in resource-constrained HIV clinics and or leading to referral burden. Our performance is comparable to recent SSA validations of the Berger scale^25,26^ and aligns with findings from southern india.^27^ Although those studies did not evaluate diagnostic discrimination.

Decision curve analysis clarified clinical utility: the ≥30 cut-off provided net benefit over “screen-none” across threshold probabilities up to 0.45, with maximum benefit at 0.30. In settings with severe mental health resource limitations, this threshold offers pragmatic triage toward patients at highest risk for confirmatory assessment.

### Clinical Recommendation

Use the ≥30 cut-off as a triage tool pending external validation in independent cohorts.

### Internal Validation and Generalisability

Internal validation confirmed stability: 10-fold cross-validated AUC (0.703) and bootstrap-corrected AUC (0.701) nearly matched the apparent estimate, indicating minimal overfitting. However, external validation is essential, as the same dataset derived and evaluated the cut-off. Good calibration of the multivariable model (Hosmer–Lemeshow p = 0.607) supports reliability. Generalizability is limited to urban tertiary settings; performance may differ in rural areas or lower-level facilities. The ≥30 threshold should be considered provisional pending broader validation.

### Stigma Dimensions: Unexpected Patterns

Disclosure concerns (96.14%) and public attitude concerns (56.49%) were highly prevalent, whereas personalised stigma (6.67%) and negative self-image (0.35%) were rare (see supplementary file). In multivariable analysis, public attitudes emerged as the strongest predictor, while disclosure concerns—despite highest prevalence—showed no independent association. The near-universal prevalence of disclosure concerns likely created insufficient variance for statistical discrimination; among those engaged in tertiary care (80% had disclosed), these concerns may reflect adaptive vigilance rather than acute distress.

Negative self-image showed a trend in univariable analysis but did not reach significance in multivariable. With only n=1 reporting high scores, any association remains tentative and hypothesis-generating. Longitudinal research is needed to clarify temporal relationships and whether these unexpected patterns reflect measurement limitations or genuine phenomenon.

### Sex Differences and Participation Bias

Females showed higher psychosocial vulnerability (49.8% vs. 32.1%). Male sex appeared protective in multivariable analysis, but this may reflect selection bias: reasons for non-participation (time constraints, competing responsibilities) may have preferentially excluded men. Sex-stratified refusal rates were not recorded; future studies should prospectively assess gender-related participation bias. The male protective effect should be interpreted cautiously. **Employment and Other Factors**

Unemployment showed bivariate association with vulnerability but did not persist in multivariable analysis, suggesting mediation by stigma dimensions and other variables. Scale discriminatory performance was comparable across sexes, and despite minor differences in sex-specific Youden cut-offs, the uniform ≥30 threshold is justified for clinical feasibility (supplementary tables).

### Contextual Literature

The Berger scale has been validated across 166 studies with growing evidence in low-resource settings.^26^ The findings from this study contribute to the literature because of the paucity of validated studies on HIV stigma in SSA and LMIC, where HIV is prevalent.^26^ Depression and anxiety are highly prevalent in sub-Saharan Africa: approximately 15.3% of PLWH meet criteria for major depression with 2-fold higher burden than non-HIV populations.^28^ Mental health directly impacts HIV outcomes: severe depression associates with 3.6-fold greater odds of delayed care-seeking.^29^ These findings support integrating mental health screening into routine HIV care, particularly in Nigeria—which bears one of Africa’s highest HIV burdens with persistent barriers including limited mental health infrastructure. WHO/UNAIDS guidance emphasizes that reaching 95-95-95 targets requires integrated psychosocial support; screening tools combined with lay counsellor-led interventions offer scalable approaches.^30^ However, tool implementation alone is insufficient; systemic barriers—workforce shortages, competing clinical priorities, and stigma rooted in gender and poverty—remain fundamental challenges requiring policy-level interventions.

The scale’s brevity and adequate psychometrics support potential integration into routine practice, with important caveats: (1) modest specificity requires confirmatory assessment; (2) external validation across diverse care settings is essential; (3) implementation requires addressing systemic barriers to mental health integration; and (4) this tool should complement—not replace—clinical judgment. Future implementation research should examine feasibility and equity impacts, with attention to scaling mental health integration toward UNAIDS 95-95-95 targets.

### Limitations

This study’s cross-sectional design prevents causal inference, and the findings are limited to an urban, tertiary-level, treatment-engaged cohort, so performance of the ≥30 cut-off may differ in rural or lower-level settings. We acknowledge that relying on two separate screening tool thresholds before applying the stigma scale cut-off introduces a nested-threshold structure that could potentially reduce specificity or introduce classification bias; nevertheless, this definition was retained because the primary clinical objective was sensitive case-finding rather than diagnostic precision. The same dataset was used to derive and evaluate the cut-off, making external validation essential before clinical adoption. Sex-stratified refusal rates were not recorded, introducing potential gender-related participation bias. Finally, the moderate discrimination indicates that stigma explains only part of the variation in psychosocial vulnerability, highlighting the need to assess multiple domains comprehensively in future studies.

## Conclusion

The 12-item Brief Berger Scale demonstrates acceptable performance for screening psychosocial vulnerability in urban tertiary HIV care in Nigeria, with the ≥30 cut-off offering high sensitivity and strong NPV as a rule-out tool pending external validation. Public attitudes emerged as the strongest vulnerability predictor. The near-universal prevalence of disclosure concerns yet their statistical absence warrants longitudinal investigation.

## Data Availability

The data underlying the findings presented in this manuscript are fully available without restriction. All relevant data are within the paper and its Supporting Information files. In addition, the raw data have been uploaded to a public repository to ensure open access and are accessible at https://doi.org/10.5281/zenodo.17088926.

## Acknowledgement

Not Applicable.

## Data Availability Statement

The de-identified dataset used in this study is publicly available in the Zenodo repository and can be accessed at 10.5281/zenodo.17088926.31

## Supporting Information Legends

**Supplementary Table 1.** Descriptive statistics for the 12-item Brief Berger HIV Stigma Scale.

Presents item means and standard deviations, average interitem covariance, subscale mean scores, and Cronbach’s alpha reliability coefficients for the four stigma dimensions (Personalized Stigma, Disclosure Concerns, Concerns about Public Attitudes, and Negative Self-Image).

**Supplementary Table 2.** Perceived HIV stigma subscale classifications.

Shows the frequency and percentage of participants classified as having high versus low stigma across the four stigma subscales.

**Supplementary Table 3. Confusion matrix for the** ≥**30 cut-off on the 12-item Brief Berger HIV Stigma Scale.**

Displays the number of true positives, true negatives, false positives, and false negatives when using a cut-off score of ≥30 to identify psychosocial vulnerability (n=285).

**Supplementary Table 4. Predictive values of the** ≥**30 cut-off at varying baseline prevalence levels.**

Reports positive and negative predictive values for hypothetical prevalence rates of 10% and 25%, based on the study’s sensitivity (87.5%) and specificity (49.0%).

**Supplementary Table 5. Participant characteristics by psychosocial vulnerability status.** Provides bivariate comparisons of sociodemographic and clinical variables between participants with and without psychosocial vulnerability, including p-values for each comparison.

**Supplementary Table 6. Univariable logistic regression analyses of stigma subscales.** Presents odds ratios, 95% confidence intervals, and p-values for associations between each of the four stigma subscales and psychosocial vulnerability.

