## Supplementary tables for "Establishing an empirical cut-off on the 12-item Brief Berger HIV Stigma Scale to screen psychosocial vulnerability among PLHIV in Nigeria"

The 12-item Berger HIV Stigma scale measures four subscales: Personalized Stigma, Disclosure Concerns, Concern about Public Attitude and Negative Self-Image. The overall scale reliability is strong (0.87), and participants reported varying levels of stigma across the subscales. While the average interitem covariance is moderate (0.27), the scale demonstrates good internal consistency among participants

**Supplementary Table 1. Descriptive statistics for items and subscales in the short-form version of the Berger Human Immunodeficiency Virus stigma scale**

| | **Mean item score (SD)** | **Average interitem covariance** | **Mean subscale score (SD)** | **Reliability α** |
|---|---|---|---|---|
| 12 - item Berger HIV Stigma scale | | 0.27 | 33.62 (6.74) | 0.87 |
| **Personalized Stigma** | | 0.44 | 5.38 (1.47) | 0.77 |
| Some people avoid touching me once they know I have HIV | 2.86 (0.79) | | | |
| People I care about stopped calling after learning I have HIV | 2.21 (0.96) | | | |
| I have lost friends by telling them I have HIV | 2.19 (0.95) | | | |
| **Disclosure Concerns** | | 0.25 | 9.91 (1.35) | 0.72 |
| Telling someone I have HIV is risky | 2.97 (0.87) | | | |
| I work hard to keep my HIV a secret | 3.16 (0.69) | | | |
| I am very careful who I tell that I have HIV | 3.24 (0.63) | | | |
| **Concerns about public attitudes** | | 0.27 | 7.66 (1.37) | 0.62 |
| People with HIV are treated like outcasts | 3.05 (0.79) | | | |
| Most people believe that a person who has HIV is dirty | 2.63 (1.07) | | | |
| Most people are uncomfortable around someone with HIV | 3.24 (0.72) | | | |

| | | | | |
|---|---|---|---|---|
| **Negative self-image** | | 0.41 | 4.74 (0.85) | 0.72 |
| I feel guilty because I have HIV | 2.65 (0.80) | | | |
| People's attitudes about HIV make me feel worse about myself | 2.89 (0.80) | | | |
| I feel I am not as good a person as others because I have HIV | 2.47 (1.14) | | | |

The perceived HIV stigma subscale data show that a majority of participants reported high stigma for Disclosure Concern (96.14%) and Public Attitude (56.49%). In contrast, Personalized Stigma was high for 6.67% of participants, while Negative Self-Image was high for only 0.35% (n=1), indicating that concerns about disclosure and public attitudes are more prevalent than internalized or personalized stigma

**Supplementary Table 2: Perceived Human Immunodeficiency Virus stigma subscale.**

| **Stigma Subscale** | **High**<br>**Freq (%)** | **Low**<br>**Freq (%)** |
|---|---|---|
| Personalized | 19 (6.67) | 266 (93.33) |
| Disclosure concern | 273 (96.14) | 11 (3.86) |
| Public attitude | 161 (56.49) | 124 (43.51) |
| Negative self-image | 1 (0.35) | 284 (99.65) |

Two-by-two classification table showing the performance of the 12-item Brief Berger HIV Stigma Scale using a cut-off score of ≥30 for identifying psychosocial vulnerability (n=285). At this threshold, the scale correctly identified 112 of 128 vulnerable participants (true positives) and 77 of 157 non-vulnerable participants (true negatives), yielding a sensitivity of 87.5% and specificity of 49.0%.

**Supplemententry Table 3: Confusion matrix for the 12-item Brief Berger HIV Stigma Scale at cut-off ≥30**

| | **Psychosocial Vulnerability Present (n=128)** | **Psychosocial Vulnerability Absent (n=157)** | **Total** |
|---|---|---|---|
| Stigma score ≥30 | 112 (True Positive) | 80 (False Positive) | 192 |
| Stigma score <30 | 16 (False Negative) | 77 (True Negative) | 93 |

| | Psychosocial Vulnerability Present (n=128) | Psychosocial Vulnerability Absent (n=157) | Total |
|---|---|---|---|
| Total | 128 | 157 | 285 |

At lower baseline prevalence levels (10% and 25%), the ≥30 cut-off maintains a high negative predictive value while the positive predictive value declines

**Supplementary Table 4: Predictive values of the ≥30 cut-off at lower hypothetical prevalence levels**

| Prevalence of Psychosocial Vulnerability | Positive Predictive Value (PPV) | Negative Predictive Value (NPV) |
|---|---|---|
| **10%** | 0.16 (16.0%) | 0.97 (97.2%) |
| **25%** | 0.36 (36.4%) | 0.92 (92.2%) |

*Sensitivity = 87.5%; Specificity = 49%.*

---

Higher total of 12-item Brief Berger HIV Stigma Scale, female sex, and unemployment were significantly associated with psychosocial vulnerability in bivariate analysis, whereas age, marital status, education, household size, income, disclosure status, and duration of HIV diagnosis were not (all p ≥ 0.2 except where otherwise noted).

**Supplementry Table 5: Participant Characteristics by Psychosocial Vulnerability Status**

| Variable | No Vulnerability (n=157) | Psychosocial Vulnerability (n=128) | p-value |
|---|---|---|---|
| **Total Berger HIV Stigma Score** | 30 (27–36) | 37 (33–41) | <0.001 (Wilcoxon) |
| **Age group (years)** | | | 0.60 |
| 21–30 | 11 (7.0%) | 14 (10.9%) | |
| 31–40 | 20 (12.7%) | 19 (14.8%) | |
| 41–50 | 71 (45.2%) | 53 (41.4%) | |
| ≥51 | 55 (35.0%) | 42 (32.8%) | |
| **Sex** | | | 0.007 |
| Female | 104 (66.2%) | 103 (80.5%) | |
| Male | 53 (33.8%) | 25 (19.5%) | |
| **Marital status** | | | 0.20 |
| Married/Co-habiting | 102 (65.0%) | 90 (70.3%) | |

| Variable | No Vulnerability (n=157) | Psychosocial Vulnerability (n=128) | p-value |
|---|---|---|---|
| Single/Widowed | 23 (14.6%) | 22 (17.2%) | |
| Divorced/Separated | 32 (20.4%) | 16 (12.5%) | |
| **Level of education** | | | 0.20 |
| No formal education | 13 (8.3%) | 10 (7.8%) | |
| Primary | 14 (8.9%) | 18 (14.1%) | |
| Secondary | 60 (38.2%) | 57 (44.5%) | |
| Tertiary | 70 (44.6%) | 43 (33.6%) | |
| **Number of people in household** | | | 0.30 |
| 1–2 | 34 (21.7%) | 36 (28.1%) | |
| 3–4 | 63 (40.1%) | 41 (32.0%) | |
| ≥5 | 60 (38.2%) | 51 (39.8%) | |
| **Employment status** | | | 0.029* |
| Full-time | 37 (23.6%) | 30 (23.4%) | |
| Part-time | 93 (59.2%) | 63 (49.2%) | |
| Retired | 9 (5.7%) | 4 (3.1%) | |
| Unemployed | 18 (11.5%) | 31 (24.2%) | |
| **Monthly income (NGN)** | | | 0.20 |
| <50,000 | 72 (45.9%) | 73 (57.0%) | |
| 50,000–100,000 | 64 (40.8%) | 42 (32.8%) | |
| >100,000 | 21 (13.4%) | 13 (10.2%) | |
| **HIV serostatus disclosure** | | | 0.60 |
| Yes (to ≥1 person) | 124 (79.0%) | 104 (81.2%) | |
| No | 33 (21.0%) | 24 (18.8%) | |
| **Duration living with HIV (years)** | | | 0.30 |
| <1 | 4 (2.5%) | 6 (4.7%) | |
| 1–4 | 15 (9.6%) | 7 (5.5%) | |
| ≥5 | 138 (87.9%) | 115 (89.8%) | |

*Fisher's exact test (employed because of small expected cell counts in retired category)

All four stigma dimensions were statistically significantly associated with psychosocial vulnerability in unadjusted analysis.

**Supplementry Table 6: Univariable Logistic Regression Analyses of Associations Between Berger HIV Stigma Subscales and Psychosocial Vulnerability**

| Stigma Subscale | Odds Ratio (OR) | 95% CI | p-value |
|---|---|---|---|
| Negative Self-Image | 1.81 | 1.35 – 2.45 | <0.001 |
| Concerns with Public Attitudes | 1.75 | 1.43 – 2.17 | <0.001 |

| Stigma Subscale | Odds Ratio (OR) | 95% CI | p-value |
|---|---|---|---|
| Disclosure Concerns | 1.39 | 1.16 – 1.67 | <0.001 |
| Personalised Stigma | 1.22 | 1.03 – 1.44 | 0.019 |
